# Non-steroidal anti-inflammatories for analgesia in critically ill patients: a systematic review and meta-analysis of randomized control trials

**DOI:** 10.1101/2023.01.03.23284166

**Authors:** Chen-Hsiang Ma, Kimberly B. Tworek, Janice Y. Kung, Sebastian Kilcommons, Kathleen Wheeler, Arabesque Parker, Janek Senaratne, Erika Macintyre, Wendy Sligl, Constantine J. Karvellas, Fernando G Zampieri, Demetrios Jim Kutsogiannis, John Basmaji, Kimberley Lewis, Dipayan Chaudhuri, Sameer Sharif, Oleksa G. Rewa, Bram Rochwerg, Sean M. Bagshaw, Vincent I. Lau

**Affiliations:** Faculty of Medicine and Dentistry, University of Alberta, and Alberta Health Services, Edmonton, Alberta, Canada; John W. Scott Health Sciences Library, University of Alberta, Edmonton, Alberta, Canada; Department of Anesthesia, McMaster University, Hamilton, Ontario, Canada; Department of Critical Care Medicine, Faculty of Medicine and Dentistry, University of Alberta, and Alberta Health Services, Edmonton, Alberta, Canada; Department of Medicine, Division of Critical Care, Western University, London, Ontario, Canada; Department of Health Research Methods, Evidence & Impact, McMaster University, Hamilton, Ontario, Canada; Department of Medicine, Division of Critical Care, McMaster University, Hamilton, Ontario, Canada; School of Public Health, University of Alberta, Edmonton, Alberta, Canada; Critical Care Strategic Clinical Network, Alberta Health Services, Edmonton, Alberta, Canada

**Keywords:** NSAID, pain, Intensive care unit, opiates, meta-analysis, ketorolac

## Abstract

**Purpose:** While opioids are part of usual care for analgesia in the intensive care unit (ICU), there are concerns regarding excess use. This is a systematic review of non-steroidal anti-inflammatories (NSAIDs) use in critically ill adult patients.

**Methods:** We conducted a systematic search of MEDLINE, EMBASE, CINAHL, and Cochrane Library. We included randomized control trials (RCTs) comparing NSAIDs alone or as an adjunct to opioids for analgesia. The primary outcome was opioid utilization. We reported mean difference for continuous outcomes and relative risk for dichotomous outcomes with 95% confidence intervals (CIs). We evaluated study risk of bias using the Cochrane risk of bias tool and evidence certainty using GRADE.

**Results:** We included 15 RCTs (n=1621 patients). Adjunctive NSAID therapy to opioids reduced 24-hour oral morphine equivalent consumption by 21.4mg (95% CI: 11.8-31.0mg reduction, high certainty) and probably reduced pain scores (measured by visual analogue scale) by -6.1mm (95% CI: -12.2 to +0.1, moderate certainty). Adjunctive NSAIDs probably had no impact on duration of mechanical ventilation (-1.6 hours, 95% CI: -0.4 to -2.7 hours, moderate certainty) and may have no impact on ICU length of stay (-2.1 hours, 95% CI: -6.1 to +2.0 hours, low certainty). Variability in reporting of adverse outcomes (e.g. gastrointestinal bleeding, acute kidney injury) precluded their meta-analysis.

**Conclusion:** In critically ill adult patients, NSAIDs reduced opioid use, probably reduced pain scores, but were uncertain for duration of mechanical ventilation or ICU length of stay. Further research is required to characterize the prevalence of NSAID-related adverse outcomes.

**Take-Home Message:** In this systematic review and meta-analysis of 15 randomized control trials that included 1621 critically ill adult patients, the addition of non-steroidal anti-inflammatories to an opioid analgesic strategy reduced 24-hour opioid use and modestly reduced pain with no impact on duration of mechanical ventilation or ICU length of stay.

## Introduction

Opioids are liberally administered in intensive care units (ICU) as part of analgesic and sedation regimens [1]. However, the prolonged opioid exposure can lead to patients developing tolerance, physical dependence, and withdrawal if abruptly discontinued [2]. These effects may carry over even after discharge from the hospital. A retrospective study of opiate-naïve ICU patients in Canada found that 20% of patients were prescribed opioids following hospital discharge, and 4% continued to use opioids after 12 months [3, 4]. In the United States, 4.1% of patients admitted to the ICU post-operatively developed new persistent opioid use [5]. Excess opioid prescription at discharge from hospital increases the risk of opioids available for inappropriate use. Even in places where opioid crisis is less pronounced (Europe and South America), concerns remain regarding the role of opioids in delirium, respiratory depression, and ileus [2, 6–9]. Thus, a clear need for alternative adjunctive analgesics (using a multi-modal approach) to reduce opioid use in the ICU for pain control is required [1].

Non-steroidal anti-inflammatory drugs (NSAIDs) are one the most well-known and prescribed over-the-counter medication classes and are effective anti-inflammatory, anti-pyretic, and analgesic agents [10]. NSAIDs such as ketorolac have been used extensively in emergency medicine and for post-operative analgesia [11–14]. NSAIDs function by inhibiting cyclooxygenase enzymes, which prevents the synthesis of prostaglandin, thromboxane, and prostacyclin [15]. Unfortunately, prostaglandin is also implicated in protecting gastric mucosa and renal hemodynamics, contributing to adverse effects of NSAID use [16]. These adverse events are well documented in the literature, but their prevalence in the ICU setting remains unclear.

The Society of Critical Care Medicine (SCCM) guidelines for the Prevention and Management of Pain, Agitation/Sedation, Delirium, Immobility, and Sleep Disruption (PADIS) from 2018 stated a weak recommendation against the use of NSAIDs in the critical care setting, citing only minor reduction in opioid use and risks of potential adverse outcomes such as kidney injury and gastrointestinal bleeding [1]. Systematic reviews published since then have suggested that the opioid-sparing effect of NSAIDs may be understated while incidence of adverse outcomes remain uncertain [17]. However, concerns remain surrounding the small number of studies analyzed as well as the inconsistent inclusion of trials in pooled analysis across the PADIS guideline and recent reviews [1, 17, 18]. Furthermore, there is evidence that lower dose NSAIDs may be beneficial for specific patient populations (e.g., emergency department, post-surgical patients: orthopedic, spinal, cardiac, abdominal, obstetrical) [13, 14, 19–24]. However, the evidence for opioid-sparing and analgesic effects of NSAIDS in critically ill populations remains uncertain.

To address this, we conducted a comprehensive and updated systematic review and meta-analysis of the available evidence on NSAID use in the critically ill adult population.

## Methods

This review followed the Preferred Reporting Items for Systematic reviews and Meta-Analyses (PRISMA) guidelines and was registered with the International Prospective Register of Systematic Reviews (PROSPERO: CRD42022332635) on May 26, 2022 [25]. The completed PRISMA checklist is included in Supplementary Table 1.

### Search Strategy

We developed the search strategy with the assistance of a medical librarian (JYK) and the strategy underwent peer review before search translation [26]. We conducted a systematic search in Ovid Medical Literature Analysis and Retrieval System Online (MEDLINE), Ovid Excerpta Medica database (EMBASE), Cumulative Index to Nursing and Allied Health Literature (CINAHL), and Cochrane Library (via Wiley) on May 29, 2022. In addition, we also searched trial registries (e.g., ClinicalTrials.gov), Google Scholar, and bibliographies from included studies as well as relevant systematic and narrative reviews. The search did not restrict results based on publication type or language. Search strategies for each database are listed in in Supplementary Table 2.

### Study Selection

Study selection was made independently and in duplicate by two investigators (CHM, KT) using Covidence systematic review software (Veritas Health Innovations, Melbourne, Australia). Titles and abstracts were screened for study design, population, and intervention. Any study that was identified as potentially eligible at this first stage was advanced to full-text review for assessment of eligibility. We resolved disagreements using a third party (VL), if needed. We recorded reasons for exclusion at the full-text review stage only.

We included randomized control trials (RCTs) that compared NSAIDs as adjunctive systemic analgesia to opioids alone in the adult critical care setting. Critical care patients included all medical, surgical, and trauma patients admitted to the intensive care unit. The intervention group consisted of NSAIDs alone or as an adjunct to opioids. Opioid dosing could be either scheduled, through patient-controlled analgesia (PCA), or administered as needed (PRN). We excluded observational cohort studies, retrospective analyses, and non-peer-reviewed research. We also excluded studies that looked at pre- or perioperative interventions or lacked opioid-only control/placebo groups.

The primary outcome was opioid utilization, which we standardized to oral morphine equivalent (OME) using published guidelines [27]. We included the following secondary outcomes: differences in pain scores, duration of mechanical ventilation, ICU and hospital length of stay, delirium, and mortality (ICU and hospital). If multiple time points were reported for opioid utilization and pain scores, we meta-analyzed the time point with the most data available. We also included safety outcomes such as incidence of adverse events (e.g., bleeding, renal dysfunction, constipation), and longer-term psychological effects (e.g., chronic pain, post-traumatic stress disorder) defined by individual study authors.

### Data Extraction

We completed data extraction independently and in duplicate by two investigators (CHM, KT) using pre-defined abstraction forms. A third reviewer resolved any discrepancies (VL). We recorded study characteristics, patient demographics, intervention details and outcomes of interest. We requested data from authors of studies with missing results, if applicable. We also extrapolated outcomes that were only presented graphically using a web plot digitizer [28].

### Risk of Bias and GRADE Assessment

We assessed risk of bias using the modified Cochrane Collaboration risk of bias tool in the following domains: sequence generation, allocation concealment, participant/investigator blinding, selective reporting, outcome blinding, addressing incomplete data, and other biases [29]. Each domain was assigned a low, probably low, probably high, or high risk of bias. We determined the overall risk of bias by taking the highest risk score in any individual domain. We assessed the overall certainty of the evidence using the Grading of Recommendations, Assessment, Development, and Evaluation (GRADE) framework [30]. We used the narrative summaries as endorsed by GRADE (moderate certainty is *probably*, low certainty is *may*, and very low certainty is *uncertain*) to describe the effect size and certainty of evidence [31]. Disagreements within the ROB and GRADE assessment was resolved by discussion and consensus.

### Data Analysis

We conducted meta-analysis using Review Manager (RevMan) software (The Cochrane Collaboration, version 5.4., Copenhagen, Denmark) using the DerSimonian and Laird random effects model to pool effect sizes for all outcomes of interest [32]. We calculated the relative risk ratio for dichotomous outcomes and the mean difference for continuous outcomes, with corresponding 95% confidence intervals. When necessary, we converted medians to mean and standard deviation using methods suggested by the Cochrane Collaboration [33].

We assessed heterogeneity using the I^2^ statistic, the X^2^ test for homogeneity, and visual inspection of the forest plots. We considered directionality, the I^2^ value, where >50% may suggest substantial heterogeneity, and perceived heterogeneity in deciding when to downgrade the certainty of the evidence for inconsistency [34]. Although we had planned to produce funnel plots and perform Egger’s weighted regression plot analysis to assess for small study effects, none of the included outcomes had sufficient included trials (at least 10 studies) to allow for this analysis.

### Subgroup analyses

Several *a priori* subgroup analyses were considered (with hypothesized direct of effect) [35]:

1. Ketorolac versus other NSAIDs (ketorolac use would result in greater opioid reduction compared to other NSAIDs);
2. Younger (age < 55) versus elderly (age ≥ 55) (NSAID-related adverse outcomes would be less in the younger patient population);
3. Higher Acute Physiology and Chronic Health Evaluation (APACHE) scores (≥25) versus lower APACHE scores (<25) (NSAIDs would be more beneficial in patients with lower APACHE scores).
4. High versus low risk of bias studies (high risk of biases would favour NSAID use)

## Results

### Search results and study characteristics

We identified 2764 citations, reviewed 73 full-text manuscripts, and included 15 randomized controlled trials in the meta-analysis [23, 36–49] (Fig 1). Table 1 provides further details on the demographics and baseline characteristics of included trials. The meta-analysis included 1621 patients with an overall mean age of 58 ± 11.3 years, 23% of which were female. The indication for ICU admission in all trials was post-operative monitoring for elective surgeries. Cardiac surgeries accounted for 12 studies, 11 of which were post-coronary artery bypass grafts. Other studies included post-spinal fusion surgery [38], gastrectomy [39], major abdominal surgery [23], and hepatectomy [48]. NSAIDs used in the trials included non-selective (diclofenac, ketoprofen, ketorolac, indomethacin) and COX-2 selective (parecoxib, valdecoxib, etodolac) inhibitors. Of note, Hynninen *et al*. compared three different NSAIDs with a placebo control group [42]. Morphine was the most common opioid used in the trials (9 out of 15 trials), but also included other opiates, such as fentanyl, tramadol, buprenorphine, codeine, oxycodone, and piritramide.

**Fig. 1.**
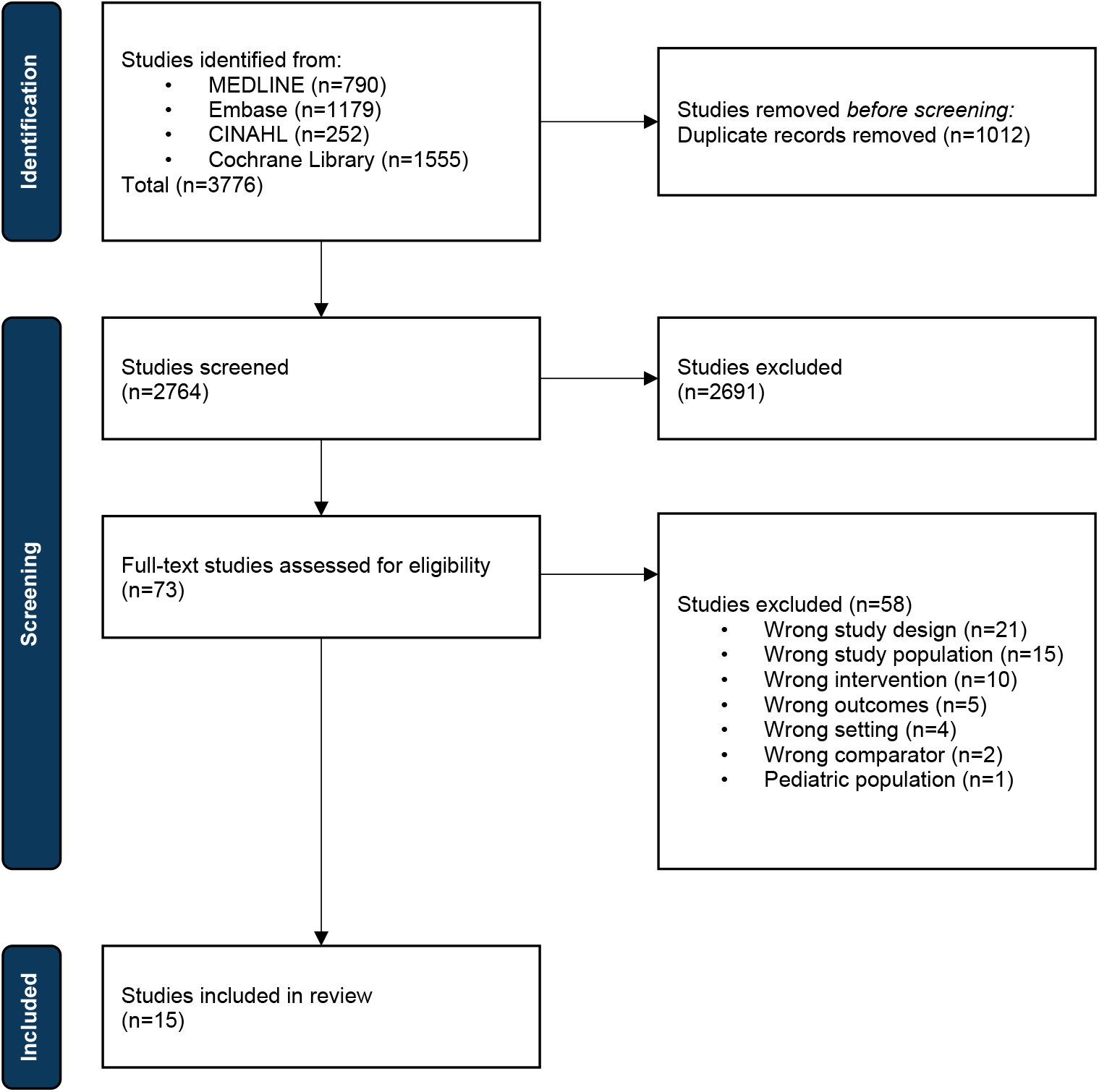
Flow chart of study selection. MEDLINE: Medical Literature Analysis and Retrieval System Online, CINAHL: Cumulative Index to Nursing and Allied Health Literature

**Table 1.** Characteristics of included studies. Data reported as mean ± sd. ICU: intensive care unit, NSAID: non-steroidal anti-inflammatory drugs, CABG: coronary artery bypass graft. Risk of bias: LR: low risk, PLR: probably low risk, PHR: probably high risk, HR: high risk

| Study | Mean Age | % Female | Sample Size | ICU Admission | NSAID Adjuvant | Opioid | Duration Of Intervention | Outcomes Analyzed | Risk Of Bias |
| --- | --- | --- | --- | --- | --- | --- | --- | --- | --- |
| **Arslan 2018** | 55.7±9.7 | 16% | 200 | Surgical-CABG | Diclofenac | Fentanyl, Tramadol | 24h | Ventilation duration, ICU length of stay | Low risk |
| **Aubrun, 2000** | 48.5±14.1 | 52% | 50 | Surgical-Spinal Fusion | Ketoprofen | Morphine | 24h | Opioid requirements, pain score | Low risk |
| **Bameshki, 2015** | 65.3±11.3 | 30% | 90 | Surgical-Gastrectomy | Diclofenac | Morphine | 24h | Opioid requirements | Low risk |
| **Barilaro, 2001** | 57.1±10.2 | 22% | 60 | Surgical-Cardiac | Ketorolac | Tramadol, Morphine | 24h | Ventilation duration | Probably high risk |
| **Fayaz, 2004** | 62.7±8.4 | 40% | 54 | Surgical-CABG | Diclofenac | Morphine | 24h | Opioid requirements, ventilation duration, post-op bleeding | High risk |
| **Hynninen, 2000** | 57.9±8.7 | 18% | 114 | Surgical-CABG | Diclofenac, Ketoprofen, Indomethacin | Morphine, Codeine, Oxycodone | 24h | Opioid requirements, post-op bleeding | Low risk |
| **Imantalab, 2014** | 20.8% (<50)<br>50.8% (50-60)<br>28.3% (>60) | 48% | 120 | Surgical-CABG | Diclofenac | Morphine | 24h | Pain score | Probably high risk |
| **Osojnik, 2020** | 66.6±8.6 | 21% | 72 | Surgical-CABG | Diclofenac | Pirtramide | 20h | Ventilation duration, ICU length of stay | Probably low risk |
| **Khalil, 2006** | 57.7±8.0 | - | 40 | Surgical-CABG | Parexocib | Morphine | 24h | Ventilation duration, post-op bleeding | Probably high risk |
| **Koizuka, 2004** | 63.6±7.4 | 23% | 26 | Surgical-CABG | Etodolac | Buprenorphine | 24h | Pain score, ventilation duration, post-op bleeding | Probably high risk |
| **Maddali, 2006** | 56.2±8.6 | 28% | 176 | Surgical-CABG | Diclofenac | Fentanyl | Until discharge from post-surgical care unit | Opioid requirements, ventilation duration, ICU length of stay | High risk |
| **Oberhofer, 2005** | 64.5±10.0 | 42% | 43 | Surgical-Abdomen | Ketoprofen | Fentanyl, Tramadol | 24h | Opioid requirements | Probably high risk |
| **Ott, 2003** | 60.6±8.1 | 13% | 462 | Surgical-CABG | Parecoxib, Valdexocib | Morphine, Codeine | 14 days | - | High risk |
| **Rapanos, 1999** | 60.9±9.5 | 21% | 57 | Surgical-CABG | Indomethacin | Morphine | 24h | Opioid requirements, pain scores, ventilation duration, post-op bleeding | Low risk |
| **Yassen, 2012** | 29.2±5.1 | 12% | 57 | Surgical-Hepatectomy | Ketorolac | Fentanyl | 48h | Opioid requirements, pain scores, ICU length of stay, post-op bleeding | Low risk |

### Risk of bias assessment

Six out of the 15 trials were deemed to have a low risk of bias [37–39, 42, 48, 49] (Table 1). Five trials had inadequate sequence generation [23, 36, 40, 44, 47], three had inadequate allocation concealment [23, 40, 47], and three trials had concerns with blinding of participants/study personnel [40, 43, 46] (Supplementary Table 3). One trial had incomplete blinding of outcome assessment [47], another did not sufficiently address missing outcome data [41], and another had concerns with selective reporting [45].

### Outcomes

Figure 2 summarizes the findings for each outcome, including the forest plot and GRADE certainty. We have also included the reasoning for rating down evidence in the GRADE analysis in Supplementary Table 4.

**Fig. 2.**
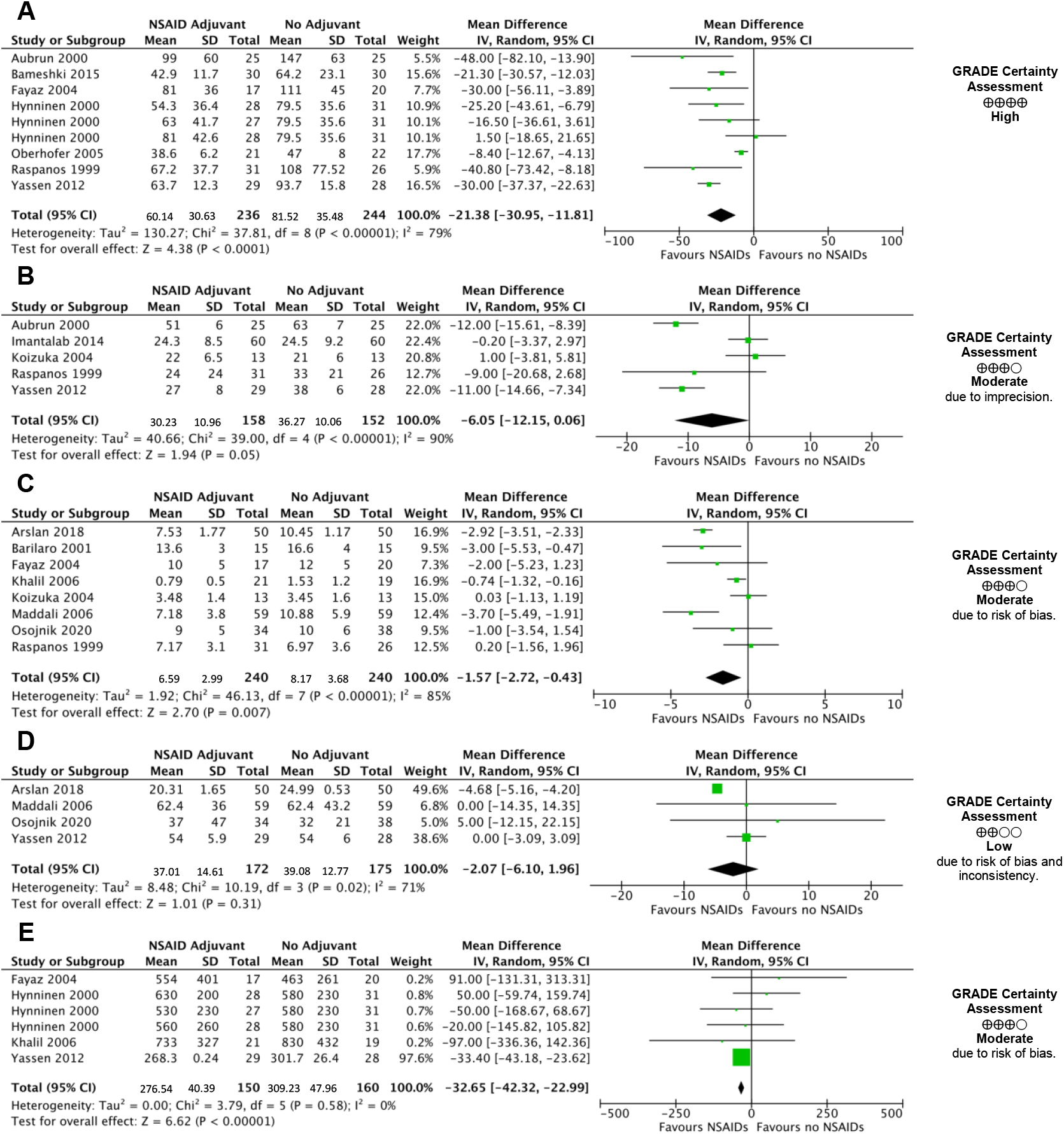
Forest plot of **A** opioid consumption in oral morphine equivalents, **B** pain scores in visual analogue scale, **C** duration of mechanical ventilation, **D** intensive care unit length of stay, and **E** bleeding

Addition of an NSAID as adjunctive analgesia reduced 24-hour oral morphine equivalent (OME) utilization by -21.4 mg (95% CI: -11.8 to -31.0mg reduction, high certainty). It also probably reduced pain measured by the visual analogue scale (VAS) by -6.1 mm (95% CI: -12.2 to +0.1, moderate certainty) at 24 hours. There was probably no impact on mechanical ventilation with the NSAID group (−1.6 hours, 95% CI: -0.4 to -2.7, moderate certainty) and NSAIDs may not have an impact on ICU length of stay (−2.1 hours, 95% CI: -6.1 to +2.0 hours, low certainty). Hospital length of stay was not reported in any of the included trials.

We were unable to complete *a priori* subgroup analyses for ketorolac versus other NSAIDs, younger age versus elderly, and higher versus lower APACHE scores due to insufficient data. Subgroup analysis by risk of bias of individual studies, shown in Supplementary Figure 1, shows no evidence of effect modification by risk of bias.

### Adverse outcomes

For the outcome of bleeding, four trials (n=265) examining blood loss after 24 hours showed that adjunctive use of NSAIDs probably did not meaningfully impact blood loss (−32.7 mL, 95% CI: -23.0 to -42.3 mL, moderate certainty) compared to opioid analgesia alone (Figure 2). Pooled estimates also show that NSAIDs have an uncertain effect on nausea and vomiting (relative risk [RR]: 0.93, 95% CI: 0.68 to 1.28, p=0.66, very low certainty) (Supplementary Figure 4). Furthermore, qualitative assessment of adverse outcomes that were not amenable to pooling did not suggest an increased risk of complications such as gastrointestinal (GI) bleeding (Table 2).

**Table 2.** Qualitative description of adverse outcomes N/A: not applicable, no significance was recorded, NS: not significant

|  | NSAID Adjuvant | No Adjuvant | Significance |
| --- | --- | --- | --- |
| **Barilaro, 2001** | ***n=15*** | ***n=15*** |  |
| ST segment elevation >0.1 mm | 1 (7%) | 1 (7%) | N/A |
| Contraction of diuresis | 2 (13%) | 0 | N/A |
| **Hynninen, 2000** | ***n=83*** | ***n=31*** |  |
| ≥20% increase in creatinine | 5 (6%) | 1 (3%) | NS |
| **Khalil, 2006** | ***n=21*** | ***n=19*** |  |
| Oliguria requiring diuretics | 16 (76%) | 9 (47%) | p=0.04 |
| Hypotension requiring inotrope support | 7 (33%) | 8 (42%) | N/A |
| **Ott, 2003** | ***n=311*** | ***n=151*** |  |
| Myocardial infarct | 5 (2%) | 1 (1%) | p=0.7 |
| Gastrointestinal bleed | 3 (1%) | 0 | p=0.6 |
| Constipation | 116 (37%) | 56 (37%) | p>1 |
| Death | 4 (1%) | 0 | p=0.3 |
| Oliguria | 15 (10%) | 45 (15%) | p=0.2 |

For the outcome of AKI defined per individual study protocol, Oberhofer *et al*. and Rapanos *et al*. described no AKI events in either group [23, 49]. Of the remaining four papers that reported on AKI, two reported no difference in the occurrence of AKI [42, 47] and one did not report statistical significance of difference in oliguria incidence [40]. Khalil *et al*. showed a statistically significant increase in oliguria treated with diuretics for patients allocated to the NSAID group compared with opioid only group [44]. However, none of the patients’ AKI progressed to being treated with renal replacement therapy [44]. Delirium was not assessed in any of the included trials.

## Discussion

This systematic review and meta-analysis demonstrated that adjunctive NSAID analgesia, compared to opioids alone, reduced 24-hour opioid utilization (high certainty evidence) and probably reduced pain scores at 24 hours (moderate certainty evidence). Adjunctive NSAIDs probably did not impact duration of mechanical ventilation (moderate certainty evidence) and may not have impacted ICU length of stay (low certainty evidence). For adverse outcomes, NSAIDs probably has no effect on blood loss 24 hours post-operatively (moderate certainty evidence) and has an uncertain effect on nausea or vomiting (very low certainty evidence). Other adverse outcomes were inconsistently reported, which prevented a pooled analysis, specifically acute kidney injury and gastrointestinal bleeding, two of the most well-known complications of NSAID use. However, a qualitative assessment of the studies suggested minimal differences in renal, GI bleeding, and cardiac dysfunction between the two groups.

The current PADIS 2018 guidelines stated a weak recommendation against using COX NSAIDs as an adjunct to opioid therapy [1]. The PADIS guideline was informed by a pooled analysis of two trials by Hynninen *et al*. and Oberhofer *et al*., which showed that adjunctive NSAID analgesia reduced 24-hour opioid use by 1.61 mg morphine equivalent (4.8 mg OME) with a non-significant reduction in pain scores at 24 hours [1, 23, 42]. The PADIS guideline concluded that the potential risk of harm superseded the small opioid-sparing effect of NSAIDs. Since then, a meta-analysis of four trials by Wheeler *et al*., including the study by Hynninen *et al*., showed that NSAIDs reduced 24-hour opioid use by 11.1 mg OME, more than double what was initially described in the PADIS guideline [17]. Our pooled analysis for opioid reduction involved seven RCTs and demonstrated that the addition of NSAIDs reduced total opioid use by 21.4 mg OME in 24 hours. Opioid utilization may be further reduced when we consider the well-documented synergistic analgesic effects of NSAID in addition to acetaminophen, a commonly used adjunctive analgesic in the critical care setting [1, 50, 51].

Our findings challenge the notion that NSAIDs may have only a small beneficial impact on reducing opioid use, although the clinical significance of the reduction remains unclear. Current evidence suggests that an absolute reduction of 10 mg IV morphine (30 mg OME) or a relative decrease of 40% in non-ICU post-operative patients over 24 hours is clinically significant [52, 53]. However, there is evidence to indicate that any opioid dose over 20 mg OME per day can increase the risk of future overdoses [54, 55]. Reducing opioid use from ≥50 mg OME daily to <20 mg can decrease the risk of overdose by half [55]. Gaps in evidence remain regarding clinically significant opioid reduction in critically ill patients, as they typically have higher opioid requirements and higher baseline pain secondary to pain from critical illness, invasive ventilation, and monitoring. In the ICU setting, where continuous infusions (0-2 mg/hr) of intravenous (IV) hydromorphone are commonly used, daily hydromorphone exposure can be up to 48 mg [56]. Our systematic review suggests that the role of NSAIDs in the arsenal of multimodal analgesia may be considered to achieve a clinically significant reduction in opioid use and possibly pain scores.

The perceived adverse events specific to NSAID use, including AKI, gastrointestinal bleeds and cardiovascular events, remain a significant barrier to their widespread use in the ICU. Although these risks have been extensively investigated, their prevalence in the ICU has not been well documented, since historically, NSAIDs have been avoided in critically ill patients [57]. Standard ICU clinical practices, which include close monitoring of creatinine, early fluid resuscitation, and stress ulcer prophylaxis, can reduce, or mitigate adverse outcomes from NSAID use [58, 59]. For example, studies in hospitalized patients with preserved kidney function have found that short-term NSAID use was not associated with an increased risk of AKI [60–62]. Even in cases of NSAID-induced AKI, cessation of the drug and fluid resuscitation usually led to a favourable prognosis and was not associated with progression to receipt of long-term dialysis [63, 64]. In cases of gastrointestinal bleeds, the co-administration of PPIs significantly reduces the risk of both duodenal and gastric ulcers and is recommended across various treatment guidelines [65, 66]. Although prior evidence suggested that NSAID use increased the risk of myocardial infarction and strokes, more recent evidence has indicated that adverse cardiac events are less than previously thought [12, 57, 67–69]. In essence, short-term NSAID use with consideration of patient risk factors and close monitoring can reduce the risk of adverse events. This paradigm is reflected in recent guidelines, which have suggested that IV ketorolac can be used as an analgesic adjunct in the ICU for up to 5 days [70]. This recommendation was based on level C quality evidence where expert opinion supported the recommendation but acknowledged a paucity of specific evidence.

Our review has several strengths. Firstly, this is the most comprehensive meta-analysis on NSAID use in the ICU to date. The adverse outcome for NSAID use (bleeding) was subjected to a pooled analysis for the first time. Further strengths are its inclusion of strictly randomized control trials, adherence to our pre-registered protocol, and inclusion of studies published in a non-English language.

Our review also has several limitations. The inclusion of only post-operative ICU patients with results limited to the first 24 hours reduces the generalizability of our findings (to the broader non-surgical ICU patient population) and may underestimate the true prevalence of adverse outcomes, particularly when NSAID exposure is prolonged. Our findings are also based on some low-quality trials with high risks of bias which is reflected in our GRADE analysis (Supplementary Table 4). However, the majority of studies are low risk of bias, and our subgroup analyses did not show any effect modification. This limitation was also present in previous systematic reviews, emphasizing the need for further, methodologically rigorous research investigating NSAIDs in ICU patients. While the opioid crisis rages worldwide, there remains a demand for adjunctive analgesics to reduce opioid consumption in the ICU setting, where other researchers are also looking at alternatives (e.g. ketamine, gabapentin, lidocaine, tramadol, etc.) [1, 17, 71–74]. While our meta-analysis indicates that NSAIDs reduce 24-hour opioid consumption, further research is required to characterize the adverse outcomes in a diverse cohort of ICU patients, exposed to NSAIDs for a longer duration. Lower dosage NSAIDs can also be considered to reduce the prevalence of adverse outcomes [75]. Ketorolac, available in intravenous formulations, has been shown in low doses to be as effective in pain relief compared to higher doses [13, 14, 19–22, 24, 48]. Therefore, further research involving a diverse ICU population, with longer term follow-up monitoring, and varied doses and durations of NSAIDs, is necessary to provide much-needed evidence on the suitability of NSAIDs in the ICU setting.

## Conclusion

This systematic review and meta-analysis found that adjunctive NSAIDs to an opioid analgesic regimen reduces 24-hour opioid utilization (high certainty evidence) and probably reduces pain scores at 24 hours (moderate certainty evidence). There was also no increased signal for harm with NSAIDs in duration of mechanical ventilation, ICU length of stay, and adverse.

## Data Availability

All data produced in the present work are contained in the manuscript.

## Acknowledgements

The authors would like to thank Alla Iansavitchene, London Health Sciences Centre, Health Sciences Library, for peer reviewing the MEDLINE search strategy.

