## Supplementary Materials for "Non-steroidal anti-inflammatories for analgesia in critically ill patients: a systematic review and meta-analysis of randomized control trials"

Chen-Hsiang Ma, BSc[1]

Kimberly B. Tworek, BScN[1]

Janice Y. Kung, MLIS[2]

Sebastian Kilcommons, BSc[1]

Kathleen Wheeler, MD[3]

Arabesque Parker, MD, MSc[4]

Janek Senaratne, MD, MSc[4]

Erika Macintyre, MD[4]

Wendy Sligl, MD, MSc[4]

Constantine J. Karvellas, MD, MSc[4]

Fernando G Zampieri, MD, PhD[4]

Demetrios Jim Kutsogiannis, MD, MSc [4]

John Basmaji, MD[5]

Kimberley Lewis, MD, MSc [6,7]

Dipayan Chaudhuri, MD, MSc[6,7]

Sameer Sharif, MD[7]

Oleksa G. Rewa, MD, MSc[4,8]

Bram Rochwerg, MD, MSc[6,7]

Sean M. Bagshaw, MD, MSc[4,8,9]

Vincent I. Lau, MD, MSc[4]

1. Faculty of Medicine and Dentistry, University of Alberta, and Alberta Health Services, Edmonton, Alberta, Canada.
2. John W. Scott Health Sciences Library, University of Alberta, Edmonton, Alberta, Canada.
3. Department of Anesthesia, McMaster University, Hamilton, Ontario, Canada.
4. Department of Critical Care Medicine, Faculty of Medicine and Dentistry, University of Alberta, and Alberta Health Services, Edmonton, Alberta, Canada.
5. Department of Medicine, Division of Critical Care, Western University, London, Ontario, Canada.
6. Department of Health Research Methods, Evidence & Impact, McMaster University, Hamilton, Ontario, Canada.
7. Department of Medicine, Division of Critical Care, McMaster University, Hamilton, Ontario, Canada.
8. School of Public Health, University of Alberta, Edmonton, Alberta, Canada.
9. Critical Care Strategic Clinical Network, Alberta Health Services, Edmonton, Alberta, Canada

Corresponding Author: Vincent Lau*, Department of Critical Care Medicine, Faculty of Medicine and Dentistry, University of Alberta and Alberta Health Services, 8440 112 Street, Edmonton, Alberta, Canada;

**Supplementary Table 1** Preferred Reporting Items for Systematic reviews and Meta-Analyses guideline checklist (DEFERRED UNTIL EDITS ARE COMPLETE!)

| Section and Topic | Item # | Checklist item | Location where item is reported |
|---|---|---|---|
| **TITLE** | | | |
| Title | 1 | Identify the report as a systematic review. | 1-3 |
| **ABSTRACT** | | | |
| Abstract | 2 | See the PRISMA 2020 for Abstracts checklist. | 4, 5 |
| **INTRODUCTION** | | | |
| Rationale | 3 | Describe the rationale for the review in the context of existing knowledge. | 6, 7 |
| Objectives | 4 | Provide an explicit statement of the objective(s) or question(s) the review addresses. | 7 |
| **METHODS** | | | |
| Eligibility criteria | 5 | Specify the inclusion and exclusion criteria for the review and how studies were grouped for the syntheses. | 8 |
| Information sources | 6 | Specify all databases, registers, websites, organisations, reference lists and other sources searched or consulted to identify studies. Specify the date when each source was last searched or consulted. | 7 |
| Search strategy | 7 | Present the full search strategies for all databases, registers and websites, including any filters and limits used. | 7 |
| Selection process | 8 | Specify the methods used to decide whether a study met the inclusion criteria of the review, including how many reviewers screened each record and each report retrieved, whether they worked independently, and if applicable, details of automation tools used in the process. | 7, 8 |
| Data collection process | 9 | Specify the methods used to collect data from reports, including how many reviewers collected data from each report, whether they worked independently, any processes for obtaining or confirming data from study investigators, and if applicable, details of automation tools used in the process. | 8, 9 |
| Data items | 10a | List and define all outcomes for which data were sought. Specify whether all results that were compatible with each outcome domain in each study were sought (e.g. for all measures, time points, analyses), and if not, the methods used to decide which results to collect. | 8 |
| | 10b | List and define all other variables for which data were sought (e.g. participant and intervention characteristics, funding sources). Describe any assumptions made about any missing or unclear information. | 8, 9 |
| Study risk of bias assessment | 11 | Specify the methods used to assess risk of bias in the included studies, including details of the tool(s) used, how many reviewers assessed each study and whether they worked independently, and if applicable, details of automation tools used in the process. | 9 |
| Effect measures | 12 | Specify for each outcome the effect measure(s) (e.g. risk ratio, mean difference) used in the synthesis or presentation of results. | 9, 10 |
| Synthesis methods | 13a | Describe the processes used to decide which studies were eligible for each synthesis (e.g. tabulating the study intervention characteristics and comparing against the planned groups for each synthesis (item #5)). | 8 |
| | 13b | Describe any methods required to prepare the data for presentation or synthesis, such as handling of missing summary statistics, or data conversions. | 9 |
| | 13c | Describe any methods used to tabulate or visually display results of individual studies and syntheses. | 9, 10 |
| | 13d | Describe any methods used to synthesize results and provide a rationale for the choice(s). If meta-analysis was performed, describe the model(s), method(s) to identify the presence and extent of statistical heterogeneity, and software package(s) used. | 9 |
| | 13e | Describe any methods used to explore possible causes of heterogeneity among study results (e.g. subgroup analysis, meta-regression). | 10 |
| | 13f | Describe any sensitivity analyses conducted to assess robustness of the synthesized results. | 10 |
| Reporting bias assessment | 14 | Describe any methods used to assess risk of bias due to missing results in a synthesis (arising from reporting biases). | 9 |
| Certainty assessment | 15 | Describe any methods used to assess certainty (or confidence) in the body of evidence for an outcome. | 9 |
| **RESULTS** | | | |
| Study selection | 16a | Describe the results of the search and selection process, from the number of records identified in the search to the number of studies included in the review, ideally using a flow diagram. | 12 |
| | 16b | Cite studies that might appear to meet the inclusion criteria, but which were excluded, and explain why they were excluded. | 8 |
| Study characteristics | 17 | Cite each included study and present its characteristics. | 10, 11, 13 |

| Section and Topic | Item # | Checklist item | Location where item is reported |
|---|---|---|---|
| Risk of bias in studies | 18 | Present assessments of risk of bias for each included study. | 13 |
| Results of individual studies | 19 | For all outcomes, present, for each study: (a) summary statistics for each group (where appropriate) and (b) an effect estimate and its precision (e.g. confidence/credible interval), ideally using structured tables or plots. | 13, 15 |
| Results of syntheses | 20a | For each synthesis, briefly summarise the characteristics and risk of bias among contributing studies. | 10, 11, 14 |
| | 20b | Present results of all statistical syntheses conducted. If meta-analysis was done, present for each the summary estimate and its precision (e.g. confidence/credible interval) and measures of statistical heterogeneity. If comparing groups, describe the direction of the effect. | 14-16 |
| | 20c | Present results of all investigations of possible causes of heterogeneity among study results. | 14 |
| | 20d | Present results of all sensitivity analyses conducted to assess the robustness of the synthesized results. | N/A |
| Reporting biases | 21 | Present assessments of risk of bias due to missing results (arising from reporting biases) for each synthesis assessed. | 14 |
| Certainty of evidence | 22 | Present assessments of certainty (or confidence) in the body of evidence for each outcome assessed. | 15 |
| **DISCUSSION** | | | |
| Discussion | 23a | Provide a general interpretation of the results in the context of other evidence. | 18 |
| | 23b | Discuss any limitations of the evidence included in the review. | 20, 21 |
| | 23c | Discuss any limitations of the review processes used. | 20, 21 |
| | 23d | Discuss implications of the results for practice, policy, and future research. | 20, 21 |
| **OTHER INFORMATION** | | | |
| Registration and protocol | 24a | Provide registration information for the review, including register name and registration number, or state that the review was not registered. | 7 |
| | 24b | Indicate where the review protocol can be accessed, or state that a protocol was not prepared. | 7 |
| | 24c | Describe and explain any amendments to information provided at registration or in the protocol. | N/A |
| Support | 25 | Describe sources of financial or non-financial support for the review, and the role of the funders or sponsors in the review. | 2 |
| Competing interests | 26 | Declare any competing interests of review authors. | 2 |
| Availability of data, code and other materials | 27 | Report which of the following are publicly available and where they can be found: template data collection forms; data extracted from included studies; data used for all analyses; analytic code; any other materials used in the review. | Within supplementary document. |

**Supplementary Table 2** Database search strategy

| Database | Search Strategy |
|---|---|
| **MEDLINE**<br><br>**Ovid MEDLINE(R) ALL** 1946 to May 27, 2022 | 1. exp Anti-Inflammatory Agents, Non-Steroidal/<br>2. NSAID*.mp.<br>3. (nonsteroidal anti-inflammator* or nonsteroidal antiinflammator*).mp.<br>4. (non-steroidal anti-inflammator* or non-steroidal antiinflammator*).mp.<br>5. (celecoxib* or etoricoxib* or lumiracoxib*).mp.<br>6. diclofenac*.mp.<br>7. etodolac.mp.<br>8. (fenoprofen* or flurbiprofen*).mp.<br>9. aspirin.mp.<br>10. ibuprofen*.mp.<br>11. indomethacin*.mp.<br>12. (ketoprofen* or ketorolac* or piroxicam* or bromfenac* or oxyphenbutazone* or suprofen*).mp.<br>13. mefenamic acid.mp.<br>14. meloxicam.mp.<br>15. naproxen*.mp.<br>16. sulindac.mp.<br>17. tolmetin.mp.<br>18. or/1-17<br>19. Analgesia/<br>20. analge*.mp.<br>21. (pain adj3 (manag* or control* or reduc* or alleviat* or mitigat*)).mp.<br>22. (pain*1 adj3 (medicat* or relief*)).tw,kf.<br>23. (pain adj2 (score* or intensit* or scale*)).tw,kf.<br>24. ((visual* analog* adj3 (scale* or score*)) and pain).tw,kf.<br>25. exp Pain Management/<br>26. Pain/dt or Pain/pc or Pain, Postoperative/dt or Pain Measurement/<br>27. or/19-26<br>28. Emergency Treatment/ or Emergency Medicine/ or Emergency Medical Services/ or Emergency Service, Hospital/ or Trauma Centers/ or Triage/ or exp Evidence-Based Emergency Medicine/ or exp Emergency Nursing/ or Emergencies/ or exp Intensive Care Units/ or Critical Care/ or Critical Illness/ or emergicent*.mp. or casualty department*.mp. or ((emergenc* or ED) adj1 (room* or accident or ward or wards or unit or units or department* or physician* or doctor* or nurs* or treatment* or visit*)).mp. or (triage or critical care or critical* ill* or (trauma adj1 (cent* or care))).mp.<br>29. (intensive care or ICU).mp.<br>30. (intensive therapy unit* or high dependency unit* or coronary care unit* or mechanical ventilat* or noninvasive ventilat*).mp.<br>31. or/28-30<br>32. 18 and 27 and 31<br>33. randomized controlled trial.pt.<br>34. clinical trial.pt.<br>35. randomi?ed.ti,ab.<br>36. placebo.ti,ab. |

| | |
|---|---|
| | 37. dt.fs.<br>38. randomly.ti,ab.<br>39. trial.ti,ab.<br>40. groups.ti,ab.<br>41. or/33-40<br>42. animals/<br>43. humans/<br>44. 42 not (42 and 43)<br>45. 41 not 44<br>46. 32 and 45 |
| **Embase**<br>**Ovid Embase** 1974 to 2022 May 27 | 1. exp nonsteroid antiinflammatory agent/<br>2. NSAID*.mp.<br>3. (nonsteroidal anti-inflammator* or nonsteroidal antiinflammator*).mp.<br>4. (non-steroidal anti-inflammator* or non-steroidal antiinflammator*).mp.<br>5. (celecoxib* or etoricoxib* or lumiracoxib*).mp.<br>6. diclofenac*.mp.<br>7. etodolac.mp.<br>8. (fenoprofen* or flurbiprofen*).mp.<br>9. aspirin.mp.<br>10. ibuprofen*.mp.<br>11. indomethacin*.mp.<br>12. (ketoprofen* or ketorolac* or piroxicam* or bromfenac* or oxyphenbutazone* or suprofen*).mp.<br>13. mefenamic acid.mp.<br>14. meloxicam.mp.<br>15. naproxen*.mp.<br>16. sulindac.mp.<br>17. tolmetin.mp.<br>18. or/1-17<br>19. analgesia/<br>20. analge*.mp.<br>21. (pain adj3 (manag* or control* or reduc* or alleviat* or mitigat*)).mp.<br>22. (pain*1 adj3 (medicat* or relief*)).tw,kw.<br>23. (pain adj2 (score* or intensit* or scale*)).tw,kw.<br>24. ((visual* analog* adj3 (scale* or score*)) and pain).tw,kw.<br>25. pain/dt or pain/pc or postoperative pain/dt or pain measurement/<br>26. or/19-25<br>27. emergency treatment/ or emergency medicine/ or emergency health service/ or hospital emergency service/ or exp evidence based emergency medicine/ or exp emergency nursing/ or emergency/ or exp intensive care unit/ or intensive care/ or critical illness/ or emergicent*.mp. or casualty department*.mp. or ((emergenc* or ED) adj1 (room* or accident or ward or wards or unit or units or department* or physician* or doctor* or nurs* or treatment* or visit*)).mp. or (triage or critical care or critical* ill* or (trauma adj1 (cent* or care))).mp.<br>28. (intensive care or ICU).mp.<br>29. (intensive therapy unit* or high dependency unit* or coronary care unit* or mechanical ventilat* or noninvasive ventilat*).mp. |

| | |
|---|---|
| | 30. or/27-29<br>31. 18 and 26 and 30<br>32. Randomized controlled trial/ or Controlled clinical study/ or randomization/ or intermethod comparison/ or double blind procedure/ or human experiment/<br>33. (random$ or placebo or (open adj label) or ((double or single or doubly or singly) adj (blind or blinded or blindly)) or parallel group$1 or crossover or cross over or ((assign$ or match or matched or allocation) adj5 (alternate or group$1 or intervention$1 or patient$1 or subject$1 or participant$1)) or assigned or allocated or (controlled adj7 (study or design or trial)) or volunteer or volunteers).ti,ab.<br>34. (compare or compared or comparison or trial).ti.<br>35. ((evaluated or evaluate or evaluating or assessed or assess) and (compare or compared or comparing or comparison)).ab.<br>36. or/32-35<br>37. (random$ adj sampl$ adj7 (cross section$ or questionnaire$1 or survey$ or database$1)).ti,ab. not (comparative study/ or controlled study/ or randomi?ed controlled.ti,ab. or randomly assigned.ti,ab.)<br>38. Cross-sectional study/ not (randomized controlled trial/ or controlled clinical study/ or controlled study/ or randomi?ed controlled.ti,ab. or control group$1.ti,ab.)<br>39. (((case adj control$) and random$) not randomi?ed controlled).ti,ab.<br>40. (Systematic review not (trial or study)).ti.<br>41. (nonrandom$ not random$).ti,ab.<br>42. Random field$.ti,ab.<br>43. (random cluster adj3 sampl$).ti,ab.<br>44. (review.ab. and review.pt.) not trial.ti.<br>45. "we searched".ab. and (review.ti. or review.pt.)<br>46. update review.ab.<br>47. (databases adj4 searched).ab.<br>48. (rat or rats or mouse or mice or swine or porcine or murine or sheep or lambs or pigs or piglets or rabbit or rabbits or cat or cats or dog or dogs or cattle or bovine or monkey or monkeys or trout or marmoset$1).ti. and animal experiment/<br>49. Animal experiment/ not (human experiment/ or human/)<br>50. or/37-49<br>51. 36 not 50<br>52. 31 and 51 |
| **CINAHL** | S1 (MH "Antiinflammatory Agents, Non-Steroidal+")<br>S2 NSAID*<br>S3 nonsteroidal anti-inflammator* or nonsteroidal antiinflammator*<br>S4 non-steroidal anti-inflammator* or non-steroidal antiinflammator*<br>S5 celecoxib* or etoricoxib* or lumiracoxib*<br>S6 diclofenac*<br>S7 etodolac<br>S8 fenoprofen* or flurbiprofen*<br>S9 aspirin<br>S10 ibuprofen*<br>S11 indomethacin*<br>S12 ketoprofen* or ketorolac* or piroxicam* or bromfenac* or oxyphenbutazone* or suprofen* |

| | |
|---|---|
| | S13 mefenamic acid<br>S14 meloxicam<br>S15 naproxen*<br>S16 sulindac<br>S17 tolmetin<br>S18 S1 OR S2 OR S3 OR S4 OR S5 OR S6 OR S7 OR S8 OR S9 OR S10 OR S11 OR S12 OR S13 OR S14 OR S15 OR S16 OR S17<br>S19 (MH "Analgesia+")<br>S20 analge*<br>S21 (pain N3 (manag* or control* or reduc* or alleviat* or mitigat*))<br>S22 (pain* N3 (medicat* or relief*))<br>S23 (pain N2 (score* or intensit* or scale*))<br>S24 ((visual* analog* N3 (scale* or score*)) and pain)<br>S25 (MH "Pain Management")<br>S26 (MH "Pain+/DT/PC")<br>S27 (MH "Postoperative Pain")<br>S28 (MH "Pain Measurement")<br>S29 S19 OR S20 OR S21 OR S22 OR S23 OR S24 OR S25 OR S26 OR S27 OR S28<br>S30 (MH "Emergency Service+") or (MH "Emergency Medicine") or (MH "Physicians, Emergency") OR (MH "Emergency Nurse Practitioners") or (MH "Emergency Nursing+") or (MH "Emergency Patients") or "casualty department*" or ((emergenc* or "ED") N1 (room* or accident or ward or wards or unit or units or department* or physician* or doctor* or nurs* or treatment* or visit*)) or (triage or (trauma N1 (cent* or care)))<br>S31 (MH "Intensive Care Units+")<br>S32 (MH "Critical Care+")<br>S33 (MH "Critical Illness")<br>S34 "intensive care" or ICU or "critical care" or critical* ill*<br>S35 intensive therapy unit* or high dependency unit* or coronary care unit* or mechanical ventilat* or noninvasive ventilat*<br>S36 S30 OR S31 OR S32 OR S33 OR S34 OR S35<br>S37 S18 AND S29 AND S36<br>S38 ( MH ( randomized controlled trials OR double-blind studies OR single-blind studies OR random assignment OR pretest-posttest design OR cluster sample ) OR TI ( randomised OR randomized ) OR AB random* OR TI trial OR ( (MH (sample size) AND AB (assigned OR allocated OR control)) ) OR MH ( placebos OR crossover design OR comparative studies ) OR AB ( (control W5 group) OR (cluster W3 RCT) OR PT (randomized controlled trial)) ) NOT ( ( MH animals+ OR MH (animal studies) OR TI (animal model*) ) NOT MH (human) )<br>S39 S37 AND S38 |
| **Cochrane Library**<br><br>via Wiley | #1 [mh "Anti-Inflammatory Agents, Non-Steroidal"]<br>#2 NSAID*<br>#3 nonsteroidal anti-inflammator* or nonsteroidal antiinflammator*<br>#4 non-steroidal anti-inflammator* or non-steroidal antiinflammator*<br>#5 celecoxib* or etoricoxib* or lumiracoxib*<br>#6 diclofenac*<br>#7 etodolac |

| | |
|---|---|
| | #8 fenoprofen* or flurbiprofen*<br>#9 aspirin<br>#10 ibuprofen*<br>#11 indomethacin*<br>#12 ketoprofen* or ketorolac* or piroxicam* or bromfenac* or oxyphenbutazone* or suprofen*<br>#13 mefenamic acid<br>#14 meloxicam<br>#15 naproxen*<br>#16 sulindac<br>#17 tolmetin<br>#18 {OR #1-#17}<br>#19 [mh Analgesia]<br>#20 analge*<br>#21 (pain NEAR/3 (manag* or control* or reduc* or alleviat* or mitigat*))<br>#22 (pain* NEAR/3 (medicat* or relief*))<br>#23 (pain NEAR/2 (score* or intensit* or scale*))<br>#24 ((visual* analog* NEAR/3 (scale* or score*)) and pain)<br>#25 [mh "Pain Management"]<br>#26 MeSH descriptor: [Pain] explode all trees and with qualifier(s): [drug therapy - DT, prevention & control - PC]<br>#27 MeSH descriptor: [Pain, Postoperative] explode all trees and with qualifier(s): [drug therapy - DT]<br>#28 [mh "Pain Measurement"]<br>#29 {OR #19-#28}<br>#30 [mh “emergency treatment”] or [mh “emergency medicine”] or [mh ^“emergency health service”] or [mh “evidence based emergency medicine”] or [mh “emergency nursing”] or [mh ^“emergency care”] or [mh “emergency ward”] or [mh “emergency”] or ( triage or casualty department* or "A&E" or "A and E"):ti,ab,kw or ((emergenc* or ED) near (room* or accident or ward or wards or unit or units or department* or physician* or doctor* or nurs* or treatment* or presentation or patient* or visit*)):ti,ab,kw<br>#31 [mh "Intensive Care Units"]<br>#32 [mh "Critical Care"]<br>#33 [mh "Critical Illness"]<br>#34 critical care or critical* NEXT ill*<br>#35 intensive care or ICU<br>#36 intensive therapy unit* or high dependency unit* or coronary care unit* or mechanical ventilat* or noninvasive ventilat*<br>#37 {OR #30-#36}<br>#38 #18 and #29 and #37 |
| **Google Scholar** | (NSAIDs OR nonsteroidal anti-inflammatory OR diclofenac OR ketorolac OR ibuprofen OR aspirin) AND (analgesia OR pain management) AND ("intensive care" OR ICU OR "critical care" OR "critical illness" OR "emergency department") AND (RCT OR trial) |

**Supplemental Table 3** Risk of bias assessment for randomized control studies

| Studies: | Adequate sequence generation (selection bias) | Allocation concealment (selection bias) | Blinding of participants/study personnel (performance bias) | Blinding of outcome assessment (detection bias) | Incomplete outcome data addressed (attrition bias) | Free from selective reporting (reporting bias) | Free from other bias | Overall risk of bias |
|---|---|---|---|---|---|---|---|---|
| **Arslan 2018** | LR | LR | LR | LR | LR | LR | LR | LR |
| **Aubrun 2000** | LR | LR | LR | LR | LR | LR | LR | LR |
| **Bameshki 2015** | LR | LR | LR | LR | LR | LR | LR | LR |
| **Barilaro 2001** | PHR | PHR | PHR | LR | LR | LR | LR | PHR |
| **Fayaz 2004** | LR | LR | LR | LR | HR | LR | LR | HR |
| **Hynninen 2000** | LR | LR | LR | LR | LR | LR | LR | LR |
| **Imantalab 2014** | LR | LR | PHR | LR | LR | LR | LR | PHR |
| **Osojnik 2020** | PLR | LR | LR | LR | LR | LR | LR | PLR |
| **Khalil 2006** | PHR | LR | LR | LR | LR | LR | LR | PHR |
| **Koizuka 2004** | LR | LR | LR | LR | LR | PHR | LR | PHR |
| **Maddali 2006** | LR | LR | HR | LR | LR | LR | LR | HR |
| **Oberhofer 2005** | PHR | PHR | LR | LR | LR | LR | LR | PHR |
| **Ott 2003** | PLR | PLR | LR | HR | LR | LR | LR | HR |
| **Rapanos 1999** | LR | LR | LR | LR | LR | LR | LR | LR |
| **Yassen 2012** | LR | LR | LR | LR | LR | LR | LR | LR |

| | |
|---|---|
| **Low risk** | LR |
| **Probably low risk** | PLR |
| **Probably high risk** | PHR |
| **High risk** | HR |
| **Unclear** | N/A |

**Supplemental Fig. 1** Subgroup analyses of low risk versus high risk of bias studies in **A** opioid consumption in oral morphine equivalents, **B** pain scores in visual analogue scale, **C** duration of mechanical ventilation, **D** intensive care unit length of stay, and **E** bleeding

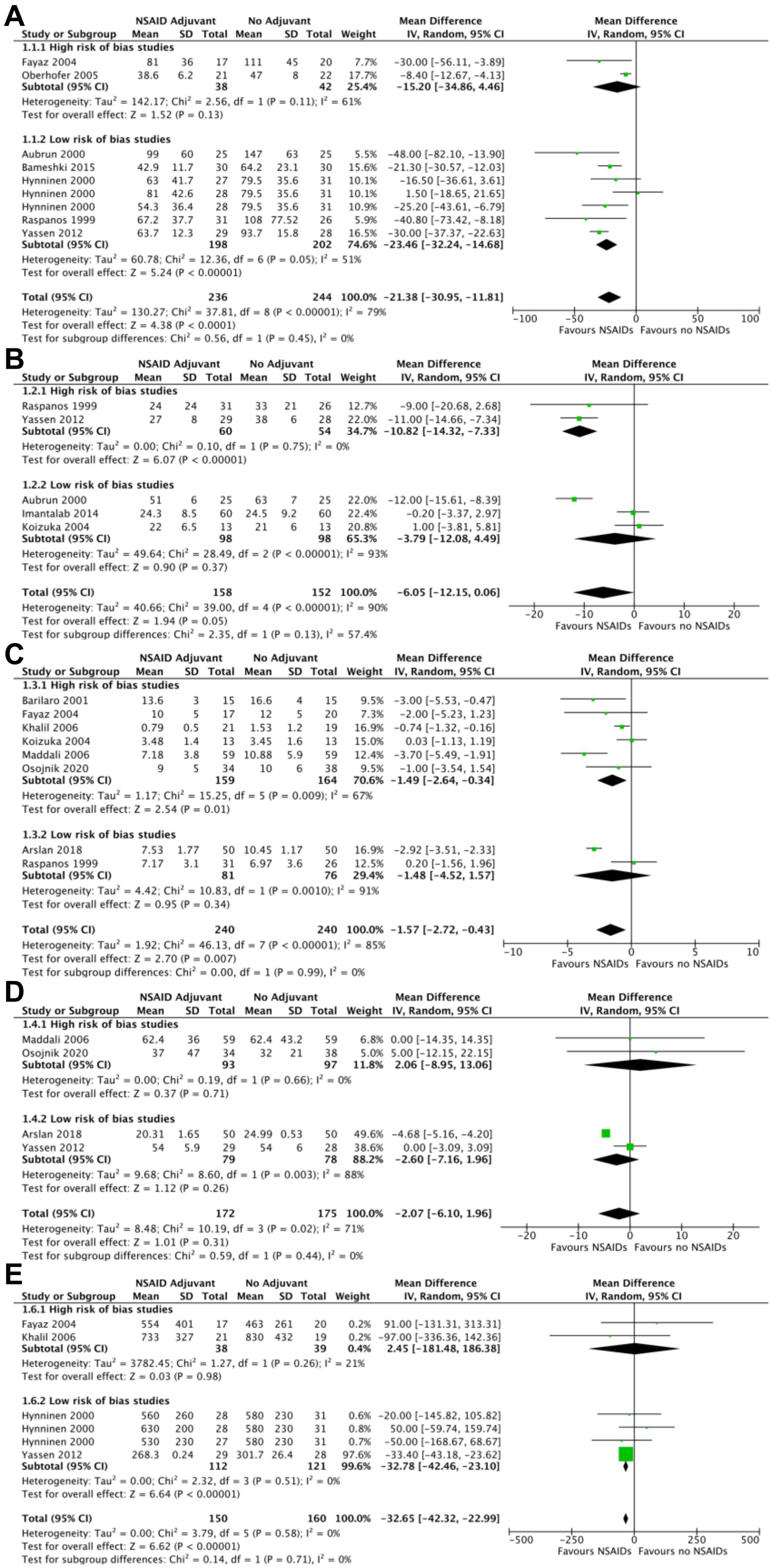

**Supplemental Table 4** Grading of Recommendations Assessment, Development and Evaluation (GRADE) outcomes

| № of studies (sample size) | Outcome mean measurement [95% CI] | | Mean difference [95% CI] | Quality assessment | | | | | | Certainty | Importance | Narrative Summary |
|---|---|---|---|---|---|---|---|---|---|---|---|---|
| | NSAID adjuvant | No NSAID | | Risk of bias | Inconsistency | Indirectness | Imprecision | Publication bias | Other considerations | | | |
| **Outcome: Reduced opiate use after 24 hours (OME, milligram)** | | | | | | | | | | | | |
| 7 RCTs (n=418) | 60.14 mg OME [56.24, 64.05] | 81.52 mg OME [77.07, 85.97] | -21.38 mg OME [-30.95, -11.81] p<0.0001 | None[a] | None[b] | None[c] | None[d] | Undetected[e] | None[f] | ⊕⊕⊕⊕ High | IMPORTANT | NSAIDs reduce opioid use in first 24 hours. |
| **Outcome: Reduced pain scores after 24 hours (VAS, millimeter)** | | | | | | | | | | | | |
| 5 RCTs (n=248) | 30.23 mm [28.52, 31.94] | 36.27 mm [34.67, 37.87] | -6.05 mm [-12.15, 0.06] p=0.05 | None[a] | None[b] | None[c] | Serious[g] | Undetected[e] | None[f] | ⊕⊕⊕◯ Moderate | IMPORTANT | NSAIDs probably reduce pain scores at 24 hours. |
| **Outcome: Duration of mechanical ventilation (hours)** | | | | | | | | | | | | |
| 8 RCTs (n=480) | 6.59 hours [6.21, 6.97] | 8.17 hours [7.70, 8.63] | -1.57 hours [-2.72, -0.43] p<0.007 | Serious[h] | None[b] | None[c] | None[i] | Undetected[e] | None[f] | ⊕⊕⊕◯ Moderate | IMPORTANT | NSAIDs probably have no impact on duration of mechanical ventilation |
| **Outcome: Intensive care unit length of stay (hours)** | | | | | | | | | | | | |
| 4 RCTs (n=347) | 37.01 hours [34.83, 39.19] | 39.08 hours [37.19, 40.97] | -2.07 hours [-6.10, 1.96] p=0.31 | Serious[h] | Serious[j] | None[e] | None[i] | Undetected[e] | None[f] | ⊕⊕◯◯ Low | IMPORTANT | NSAIDs may have no impact on ICU length of stay. |
| **Outcomes: Total bleeding after 24 hours (millilitres)** | | | | | | | | | | | | |
| 4 RCTs (n=265) | 276.54 mL [270.08, 283.01] | 309.23 mL [301.79, 316.66] | -32.65 mL [-42.32, -22.99] p<0.00001 | Serious[h] | None[k] | None[e] | None[i] | Undetected[e] | None[f] | ⊕⊕⊕◯ Moderate | IMPORTANT | NSAIDs probably have no impact on total bleeding in first 24 hours. |
| **№ of studies (sample size)** | **Anticipated absolute effects** | | **Relative effect [95% CI]** | **Quality assessment** | | | | | | **Certainty** | **Importance** | Narrative Summary |
| | **Risk with NSAID Adjuvant (n, %)** | **Risk without NSAID (n, %)** | | **Risk of bias** | **Inconsistency** | **Indirectness** | **Imprecision** | **Publication bias** | **Other considerations** | | | |
| **Outcome: Prevalence of nausea or vomiting** | | | | | | | | | | | | |
| 8 RCTs (n=543) | 52/297, 18% | 64/308, 21% | 0.93 [0.68, 1.28] p=0.66 | Serious[h] | None[k] | None[e] | Very serious[l] | Undetected[e] | None[f] | ⊕◯◯◯ Very low | LOW IMPORTANCE | NSAIDs have an uncertain effect on nausea/vomiting. |

RCT: Randomized controlled trials, CI: Confidence interval, NSAID – Non-steroidal anti-inflammatory, OME: oral morphine equivalents, VAS: visual analogue scale

**GRADE Explanations**

a. Majority of papers had low risk of bias, subgroup analysis did not suggest effect modification based on risk of bias.
b. Heterogeneity favors NSAID adjuvant and reflects variability in magnitude.
c. No indirectness since all studies compared NSAID to no NSAID adjuvant analgesia.
d. 95% CI interval range does not cross no effect and meets optimal information size.
e. No conflicts of interest in any of the papers included in the analysis. Furthermore, we reduced bias by adopting a comprehensive search strategy.
f. No upgrades for dose dependent response or magnitude of effect.
g. 95% CI crosses no effect
h. Majority of papers had increased risk of bias
i. No difference within 95% CI range.
j. High degree of heterogeneity
k. $I^2$=0%
l. 95% CI crosses no effect and insufficient optimal information size.
